# Identification of non-small cell lung cancer (NSCLC) patients who benefit from treatment beyond progression with pembrolizumab from individual lesion response dynamics

**DOI:** 10.1101/2022.05.09.22274626

**Authors:** Timothy Qi, Yanguang Cao

**Affiliations:** Division of Pharmacotherapy and Experimental Therapeutics, Eshelman School of Pharmacy, The University of North Carolina at Chapel Hill, Chapel Hill, NC 27599, USA; Lineberger Comprehensive Cancer Center, The University of North Carolina at Chapel Hill, Chapel Hill, NC 27599, USA

**Keywords:** Immunotherapy, Pembrolizumab, Virtual Clinical Trial, NSCLC, Treatment Beyond Progression

## Abstract

**Background:** Pembrolizumab is the recommended first-line treatment for non-small cell lung cancer (NSCLC) with no driver alterations and PD-L1 Tumor Proportion Score (TPS) ≥ 50%. Salvage therapies for patients who develop resistance in this setting are limited. Several retrospective studies have highlighted a subset of patients who benefit from pembrolizumab treatment beyond progression (TBP), but these results have yet to be validated in a rigorous prospective study.

**Methods:** Due to the heterogeneity of within-patient responses to pembrolizumab, many patients can experience progressive disease while some of their lesions are still shrinking. We employed nonlinear mixed-effects modeling and virtual clinical trial simulations to evaluate the risk of unconditionally switching all progressors to salvage chemotherapy and to identify the subset of patients likely to benefit from pembrolizumab TBP. By using > 25,000 radiographic lesion diameter measurements from > 500 patients, we simulated individual lesion responses to pembrolizumab, chemotherapy, and pembrolizumab treatment failure followed by pembrolizumab TBP or salvage chemotherapy. We then assessed the benefit of pembrolizumab and chemotherapy in combination and explored a potential mechanism of intrapatient drug synergy called lesion-level independent action (LLIA).

**Results:** Switching all progressors to salvage chemotherapy was suboptimal. Pembrolizumab TBP could extend progression-free survival (PFS) and control tumor burden more in nontarget progressors with fewer lesions prior to treatment initiation. Assuming LLIA but not patient-level independent action (PLIA) for combination therapy yielded longer PFS.

**Conclusions:** Pembrolizumab TBP may benefit a subset of patients with PD-L1-high, driver alteration-free NSCLC who experience nontarget progression, but prospective studies are warranted.

## INTRODUCTION

Pembrolizumab is an antibody inhibitor of programmed cell death protein 1 (PD-1) recommended for the first-line treatment of advanced PD-L1 TPS ≥ 50% (PD-L1-high) non-small cell lung cancer (NSCLC) without driver alterations [1]. However, within-patient responses to pembrolizumab are heterogeneous, with spatiotemporally dissociated patterns of response and progression frequently reported [2], [3]. This leads to situations under RECIST v1.1 where patients with nontarget progression or new metastases can be classified as having “progressive disease” (PD) regardless of target lesion response [3], [4]. As a result, patients who progress on pembrolizumab might be switched to salvage chemotherapy even while some or most of their lesions are still responding [5]. Determining the optimal salvage therapy for treatment failure in the PD-L1-high, driver alteration-free setting remains a challenge given the limited repertoire of approved therapeutics.

Several retrospective studies have found that a subset of NSCLC patients who progress under immunotherapy can still benefit from treatment beyond progression (TBP) [6]–[9]. Biomarkers of this subset have not been conclusively identified. We hypothesized that such a biomarker might be found within the intrapatient heterogeneity of individual lesion responses to pembrolizumab. This heterogeneity is well-appreciated across anatomical sites: lesions in the liver, for example, respond poorly to pembrolizumab and correlate with inferior patient-level responses in NSCLC [2]. Ethical considerations have precluded rigorous prospective studies of the relationship between TBP and lesion-level response heterogeneity, leaving biomarkers for patients likely to benefit from TBP yet undiscovered [10].

To identify the patterns of PD most likely to respond to TBP with pembrolizumab, we employed a computational approach that leveraged publicly available lesion-level growth dynamics of NSCLC treated with pembrolizumab or platinum doublet chemotherapy. Importantly, while we were able to directly obtain lesion-level growth dynamics for NSCLCs treated with chemotherapy, we needed to develop a method to infer them for NSCLCs treated with pembrolizumab due to a paucity of accessible data. These growth dynamics were specific to the lesions’ anatomical locations. We also adapted a published nontarget progression model for melanoma treated with immunotherapy to approximate nontarget progression rates under pembrolizumab [4]. Henceforth, we refer to progression caused by the appearance of new metastases or the growth of nontarget lesions collectively as “nontarget progression”.

With these data, we generated a virtual cohort of 1,000 patients with realistic distributions of baseline tumor burden across anatomical sites. After demonstrating our model could robustly recapitulate monotherapy responses to chemotherapy and pembrolizumab, we examined patients who progressed on pembrolizumab and simulated treatment with either salvage chemotherapy or pembrolizumab TBP. Strikingly, we found that progression-free survival (PFS) was indistinguishable between salvage chemotherapy and pembrolizumab TBP in patients whose original PD was caused by nontarget progression. These nontarget progressors had comparable tumor control on salvage chemotherapy and TBP and significantly fewer lesions at baseline.

We then used our model to simulate the potential benefit of combination chemotherapy and pembrolizumab. Palmer et al. demonstrated that the vast majority of clinical benefit derived from combination oncology therapies is attributable to independent drug action rather than bona fide drug synergy [11], [12]. In these analyses, only combination therapy regimens whose clinical benefit exceeded that which would be predicted from the independent action of their constituent monotherapies were accepted as potentially possessing drug synergy. We termed this framework “patient-level independent action” (PLIA) and applied it to our combination therapy simulations. Finally, we explored lesion-level independent drug action (LLIA) as a potential mechanistic framework by which drug synergy might be achieved and found it a useful supplement to PLIA.

## METHODS

### Creating a virtual cohort with anatomically distributed baseline tumor burden

Lesion-level response dynamics from [13] were obtained from Project Data Sphere (registration number: NCT00540514). The study was given ethical approval by institutional review boards at each participating center and conducted in accordance with the principles of the Declaration of Helsinki and Good Clinical Practice Guidelines of the International Conference on Harmonization. All patients provided written informed consent before study initiation. The control arm comprised 25,708 lesion diameter measurements from 524 patients with previously untreated advanced NSCLC. As driver alteration status and PD-L1 expression were unavailable in this dataset due to its time of conduct, a core assumption of our analysis was that these factors did not substantially alter the anatomical distribution of tumor burden prior to treatment initiation. We cleaned these data such that each lesion’s site corresponded to either one of seven anatomical sites reported by [2] – adrenal, bone, liver, lung, lymph node, pleural, soft tissues – or “other”. Next, target lesion diameter measurements made prior to or within 1 day after treatment initiation were designated as baseline measurements. We constructed the final bootstrapped virtual cohort (1,000 patients with 4,109 lesions) from the cleaned data by sampling and replacement.

### Obtaining lesion growth dynamics parameters under chemotherapy

Lesion-level response dynamics from [13] were segregated by anatomical site. We applied a nonlinear mixed-effects population modeling approach to estimate lesion growth dynamics parameters *f, d*, and *g* for lesions in each anatomical site [14], [15]:

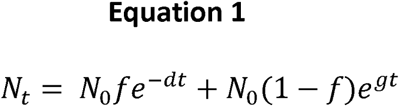

Where N_t_ represents tumor burden at time *t*, N_0_ represents baseline tumor burden, *f* represents the fraction of treatment-sensitive cells at baseline, *d* represents the death rate of treatment-sensitive cells, and *g* represents the growth rate of treatment-resistant cells. Default software recommendations in Monolix 2021 R1 [16] were used to select error models and correlation models. The best objective response (BOR) of each lesion was calculated by taking the nadir of non-baseline measurements for each patient and stored alongside its corresponding *f, d*, and *g* parameters. The end result was a “chemotherapy response matrix” containing values for *f, d, g*, and anatomical site for each lesion in [13].

### Simulating chemotherapy

All treatment simulations were conducted in MATLAB R2020b. To simulate treatment, each lesion was assigned a random vector of *f, d*, and *g* parameter values from the chemotherapy response matrix given its anatomical site. These parameters were then used with Equation 1 to simulate lesion-level response dynamics under 18 weeks of chemotherapy with radiographic assessment every 6 weeks [13]. Patient-level tumor burden was calculated by summing the diameters of all lesions within a patient at any given time. Response and PFS were calculated per RECIST v1.1 from tumor measurements at each simulated radiographic assessment. Of note, we classified PD as due to target, nontarget, or target and nontarget progression by adapting the total tumor burden-based nontarget progression model in [4] to NSCLC with data from [17]. Objective response rate (ORR) was calculated by summing the proportion of patients who achieved partial response (PR) or complete response (CR). Disease control rate (DCR) was calculated similarly and included those who achieved stable disease (SD).

### Obtaining lesion growth dynamics parameters under pembrolizumab

Patient-level tumor burden dynamics of 88 driver alteration-free NSCLC patients treated with first-line pembrolizumab monotherapy were retrospectively reported in [18]. The majority (84%) of these patients had PD-L1 TPS ≥ 50% prior to treatment initiation. Without access to the underlying data, we manually extracted the tumor burden dynamics of 77 patients with WebPlotDigitizer [19]. This provided sufficient data to use within Monolix 2021R1 to perform the same nonlinear mixed-effects population modeling approach and obtain parameter values for *f, d*, and *g*. We then calculated the BORs of patient-level tumor burden profiles by taking the nadir of non-baseline measurements for each patient. The BOR of each patient was stored in a “BOR matrix” alongside its corresponding *f, d*, and *g* parameters. We also calculated ✉ for each patient [20]:

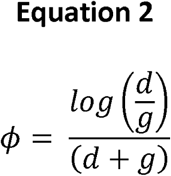

Where ⍰ is a composite lesion growth dynamics metric. Next, we extracted anatomical site-specific best responses of 480 lesions treated with anti-PD-1 monotherapy in [2] using WebPlotDigitizer [19]. Best responses for each lesion were compared against the nearest BORs in the BOR matrix and assigned their associated *f, d, g*, and ⍰ parameters. Because the lesion BORs in [2] were from a primarily pretreated population, indexing within the BOR matrix containing response parameters under first-line pembrolizumab [18] was done after adjusting for the difference in patient-level BORs between the two studies. The end result was a scaled “pembrolizumab response matrix” containing values for *f, d, g*, ⍰, and anatomical site for each lesion in [2].

### Simulating pembrolizumab monotherapy with intrapatient correlations in growth dynamics

For *n* lesions in a patient, *n* lists of site-specific ⍰ were obtained and padded by replication until all lists were of identical length. Lists were then horizontally concatenated. Values of ⍰ within each column were ranked, spiked with Gaussian noise, and reranked. An average linear correlation was calculated by performing an inverse Fisher transform on the average of all Fisher-transformed pairwise Pearson’s *ρ* between site-specific lists. In other words, this value represented the average pairwise correlation in ⍰ ranks between any two given lesions within the patient. If this average *ρ* deviated from the desired *ρ* by more than an acceptable level of tolerance, the magnitude of Gaussian noise was adjusted to increase or decrease the average pairwise lesion correlation in ⍰. Once the desired *ρ* was achieved, a random row vector from the list was sampled such that n values of ⍰ were obtained. Each lesion’s ⍰ value was used to assign it a corresponding set of *f, d*, and *g* parameter values from the pembrolizumab response matrix. For tumors located in the “other” compartment, we sampled from the pooled responses of all other sites. Based on the observation in [2] that ∼20% of patients treated with pembrolizumab exhibit highly synchronous responses, the top 20% ⍰ ranks were protected from Gaussian noise and re-ranking prior to the calculation of *ρ*.

We used *f, d*, and *g* parameters and Equation 1 to simulate 16 weeks of treatment with radiographic assessment every 8 weeks [18]. Patient-level tumor burden, ORR, DCR, and PFS were calculated as they were for chemotherapy.

### Simulating salvage chemotherapy or pembrolizumab TBP

Virtual patients were treated with pembrolizumab monotherapy for up to 16 weeks or until PD, at which point progressors were cloned in silico and given either salvage chemotherapy, pembrolizumab TBP, or a hypothetical PFS-optimized regimen. This subsequent round of therapy was maintained for 18 weeks, with radiographic assessment simulated every 6 weeks. Salvage chemotherapy was simulated by replacing the pembrolizumab-specific *f, d*, and *g* parameters of a patient’s lesions with chemotherapy-specific *f, d*, and *g* parameters.

In patients treated with PFS-optimized therapy, tumor burden was simulated pro forma under 18 weeks of either salvage chemotherapy or pembrolizumab TBP in parallel. The treatment that resulted in longer PFS was retroactively selected for the subsequent round of therapy. This was repeated 1,000 times, with the simulation yielding the median fraction of patients for whom pembrolizumab TBP yielded longer PFS than salvage chemotherapy selected for further analysis.

### Simulating combination therapy

Virtual patients were treated with pembrolizumab and chemotherapy in combination by simulating pro forma tumor burden under 18 weeks of either chemotherapy or pembrolizumab, during which two modes of independent drug action were assessed: patient-level independent action (PLIA) and lesion-level independent action (LLIA). To simulate PLIA as reported in [11], [12], the treatment that resulted in longer PFS was retroactively designated as the response for that patient. This is identical to the PFS-optimized approach mentioned previously. Under LLIA, the better outcome of either drug for each lesion was retroactively designated as the response for that individual lesion. Patient-level responses were then calculated by aggregating responses across lesions. This was repeated 1,000 times, with the simulation yielding the median proportion of patients who stood to benefit from LLIA over PLIA selected for further analysis.

## RESULTS

### Model simulations recapitulate heterogeneous clinical responses to treatment

Baseline tumor burden, lesion growth dynamics under treatment, and incidence of nontarget progression as a function of tumor burden were adapted from the literature into a tumor growth model (Figure 1A-B) [2], [4], [13], [17], [18]. These data were used to create a virtual cohort of 1,000 patients with realistic baseline tumor burden, anatomical lesion distribution, nontarget progression rate, and site-specific lesion growth dynamics under treatment. Simulated platinum doublet chemotherapy was able to recapitulate the heterogeneity and objective response rate (ORR) of patients in the control arm of [13], who received first-line carboplatin and paclitaxel (Figure 1C).

**Figure 1.**
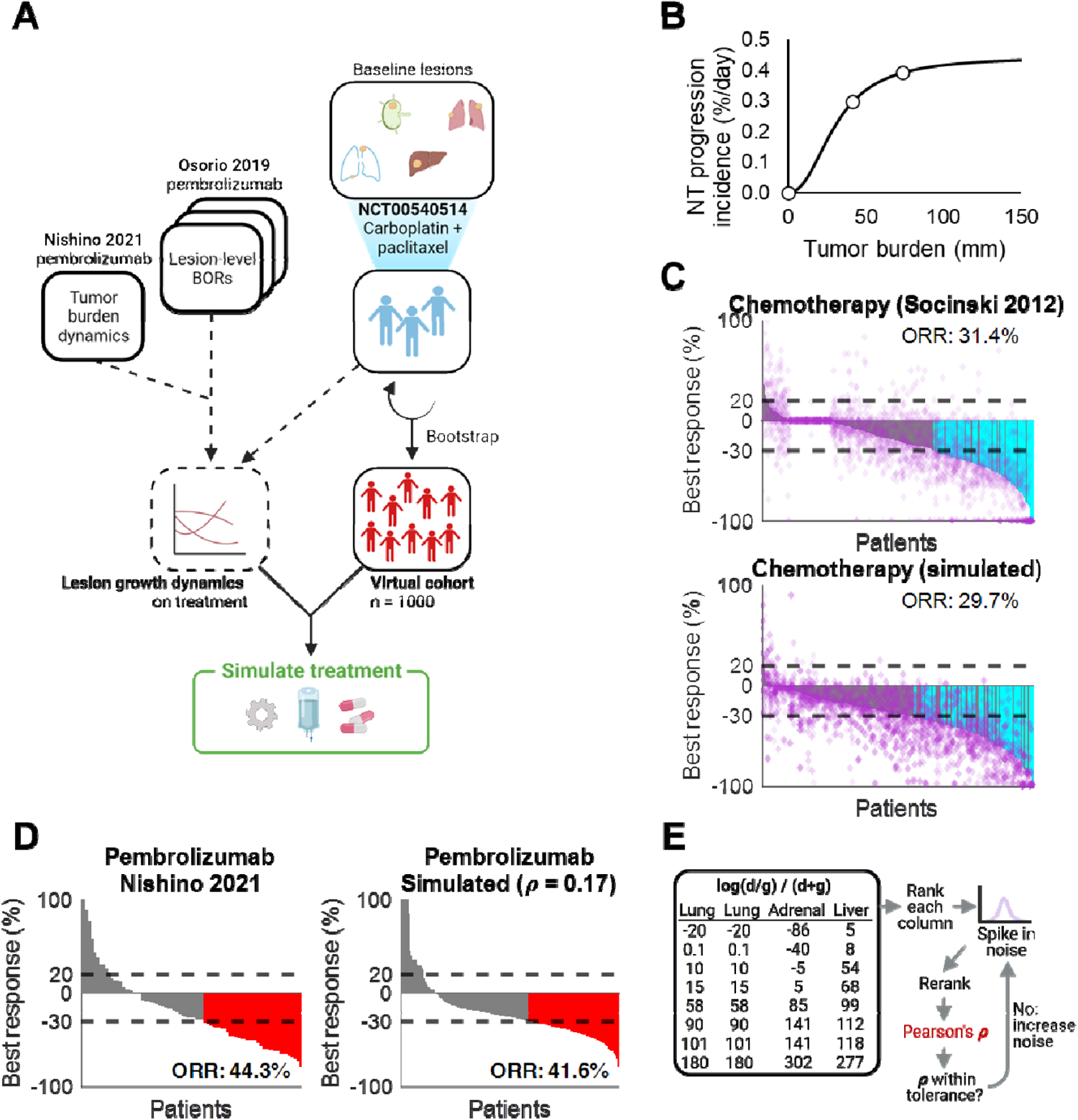
Virtual monotherapy recapitulates clinical objective responses. A. Schematic of method for generating a virtual cohort, deriving lesion growth dynamics, and simulating treatment. Data in [13] were obtained from Project DataSphere and used to bootstrap a virtual cohort of 1,000 patients. Anatomical site-specific lesion size measurements of NSCLCs in [13] were then used to fit a nonlinear mixed-effects model of lesion-level growth dynamics under platinum doublet therapy. Similarly, anatomical site-specific lesion growth dynamics were obtained from [2] and used in tandem with tumor burden dynamics from [18] to fit a nonlinear mixed-effects model for lesion growth dynamics under pembrolizumab (see Methods). B. Model for daily incidence of nontarget progression adapted from [4] with data from [17]. A Hill equation defines the relationship between total tumor burden and nontarget progression incidence. C. Monotherapy responses to platinum doublet therapy reported in [13] (top) and simulated patients (bottom). Horizontal dashed lines indicate thresholds for PD and PR; purple diamonds represent individual lesions; cyan bars indicate patients with responses and no nontarget progression; gray bars indicate patients with responses and simultaneous nontarget progression. Treatment was simulated for 18 weeks, with radiographic assessment for response every 6 weeks. D. Monotherapy responses to pembrolizumab in treatment-naïve patients digitized from [18] (left) and simulated patients (right). Horizontal dashed lines indicate thresholds for PD and PR; red bars indicate patients with PR or better based on target lesion dynamics only; *ρ* represents the average correlation of responses among lesions within each patient (see Methods). Neither individual lesion response nor nontarget progression were assessed due to lack of availability in the published dataset. E. Schematic for method of identifying the average correlation of best responses among lesions within each patient adapted from [11] (see Methods, Supplemental Figure 1).

To simulate the lesion-level growth dynamics of NSCLC treated with pembrolizumab without direct access to data, we inferred and empirically estimated the correlation in growth dynamics across lesions in each patient by simulating with correlation in response anywhere from perfect (*ρ* = 1) to nonexistent (*ρ* = 0) (Figure 1D-E; see Methods and Supplemental Figure 1). We reasoned that systemic factors such as patient immune status might mediate diverse patterns of synchronous and dissociated response that contribute to ORR. However, there was no discernible relationship between inter-lesion response correlation and ORR (Supplemental Figure 1). The *ρ* that minimized the simulated cohort’s deviance in ORR and PD from [18] was selected for further analyses. Overall, our modeling approach allowed us to create a large virtual cohort that responded realistically to simulated treatment with chemotherapy and pembrolizumab.

### Patterns of response vary with therapy and lesion location

We next explored the heterogeneity of lesion responses to therapy across anatomical sites. Virtual patients were treated for 16 to 18 weeks with periodic radiographic assessment of tumor diameters (Figure 2A). We found that first-line chemotherapy and pembrolizumab yielded comparable tumor control and PFS within this time frame, consistent with clinical observations [21] (Figure 2B-C). Furthermore, lesions in the liver and soft tissue that were at high risk of nonresponse to pembrolizumab tended to respond well to chemotherapy (Figure 2D).

**Figure 2.**
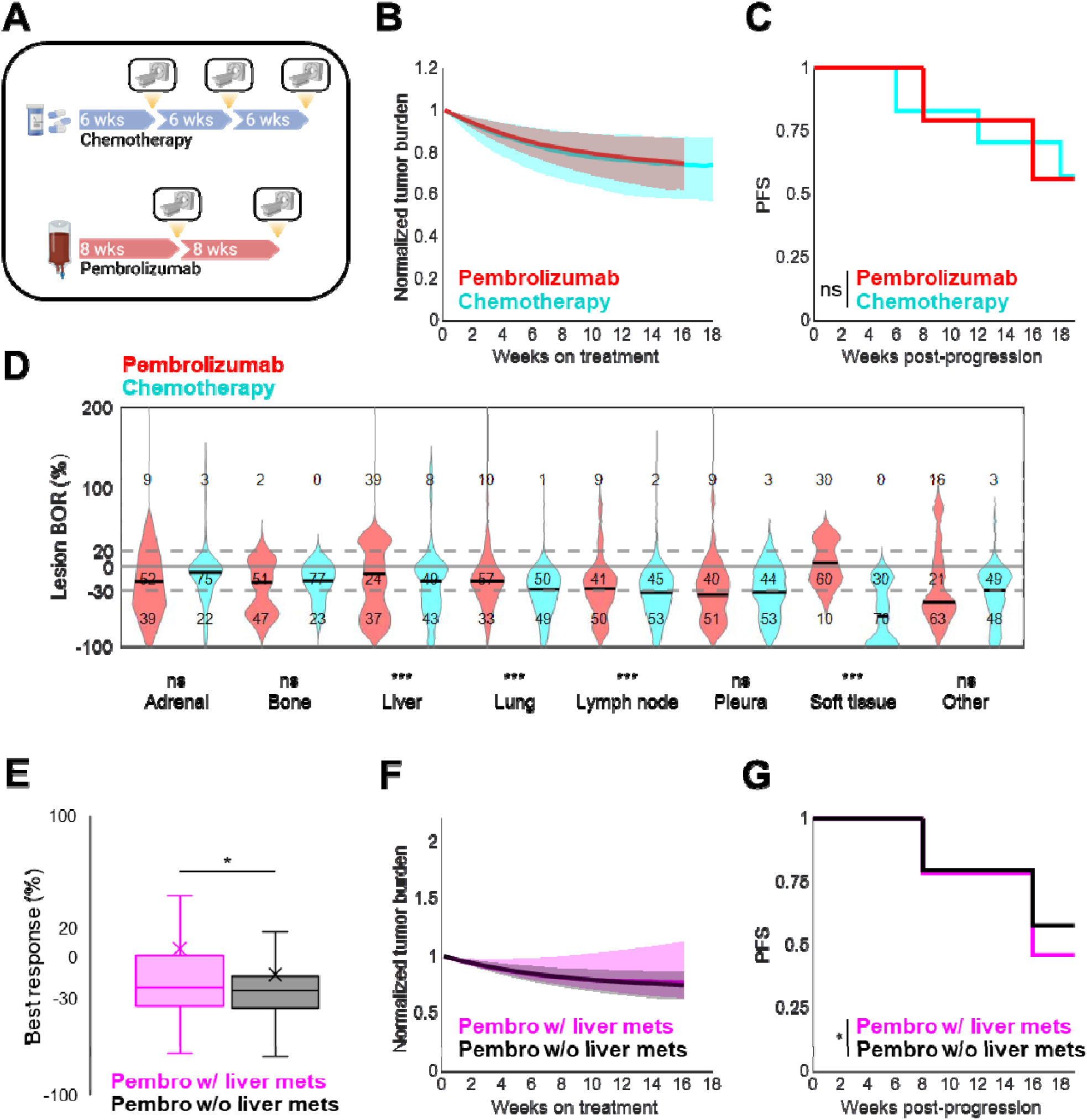
Responses to virtual monotherapy vary across anatomical sites. A. Schematic of virtual clinical trial comparing pembrolizumab versus chemotherapy. Treatment with chemotherapy was simulated for 18 weeks with radiographic assessment for response every 6 weeks. Treatment with pembrolizumab was simulated for 16 weeks with radiographic assessment for response every 8 weeks. B. Median tumor burden under chemotherapy (cyan) and pembrolizumab (red). Shaded regions represent the interquartile range at each time point. C. PFS under chemotherapy (cyan) and pembrolizumab (red). Ns, not significant (log-rank test). D. Lesion-level BORs under chemotherapy (cyan) and pembrolizumab (red) by anatomical location. Gray lines indicate cutoffs for PD and PR; solid black lines indicate medians. Ns, not significant; ***, p < 0.001 (unpaired Student’s t-test). E. Best patient-level reduction in tumor burden under pembrolizumab in patients with (pink) and without (black) baseline liver metastases. X, mean; *, p < 0.05 (unpaired Student’s t-test). F. Median tumor burden under pembrolizumab in patients with (pink) and without (black) baseline liver metastases. Shaded regions represent the interquartile range at each time point. G. PFS under pembrolizumab in patients with (pink) and without (black) baseline liver metastases). *, p < 0.05 (log-rank test).

Baseline lesion distribution across anatomical sites did not dramatically affect lesion growth dynamics (Supplemental Figure 2A-B). However, previous studies reported that liver metastases reduced the efficacy of anti-PD-1 therapy in NSCLC [2], [22]. In our virtual cohort, patients with liver metastases prior to treatment initiation also experienced shallower best responses and shorter PFS under pembrolizumab than those without (Figure 2E-G). Conversely, liver metastases conferred no difference in PFS under chemotherapy, although they did affect best responses and tumor control (Supplemental Figure 2C-E). Baseline metastases in select locations non-central nervous system locations, such as the liver, may be an important patient selection criterion for prospective clinical studies involving immunotherapy.

### Unconditionally-applied salvage chemotherapy after progression on pembrolizumab yields suboptimal progression-free survival

With successful recapitulation of lesion-level responses to monotherapy, we next conducted a pembrolizumab salvage therapy trial to identify which patients, if any, could benefit from pembrolizumab TBP (Figure 3A). Virtual patients were treated with pembrolizumab for up to 16 weeks or until PD, at which point they received pembrolizumab TBP or salvage chemotherapy for 18 additional weeks. Progressors were stratified by their initial cause of PD: target progression only, nontarget progression only, or simultaneous target and nontarget progression at the time of radiographic assessment. Progression-free survival under subsequent therapy (PFS2) was then assessed in each group.

**Figure 3.**
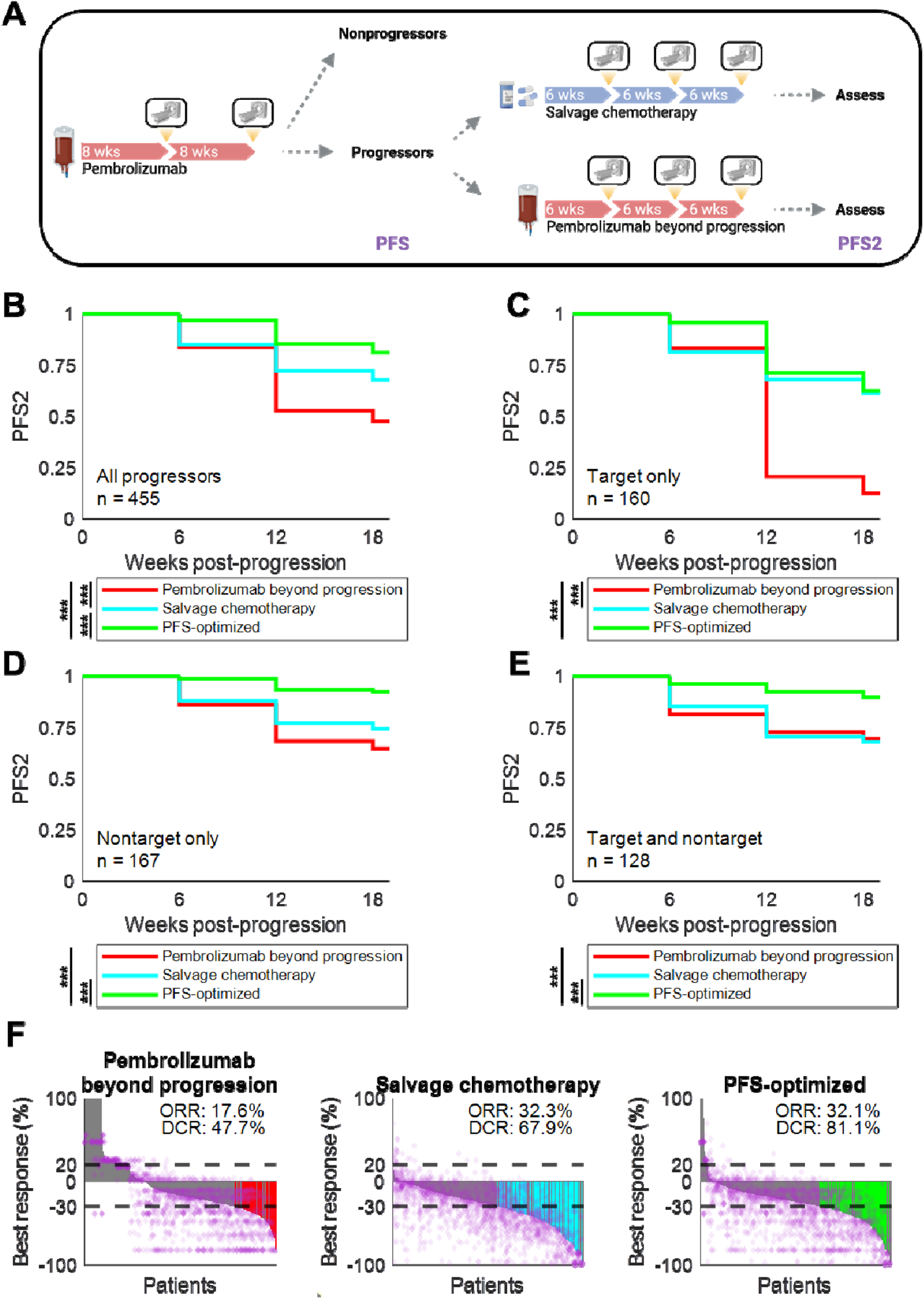
Pembrolizumab TBP can prolong PFS in nontarget progressors. A. Schematic of a virtual clinical trial comparing pembrolizumab TBP versus salvage chemotherapy. Treatment with pembrolizumab was simulated for up to 16 weeks with radiographic assessment for response every 8 week. Progressors were immediately cloned and received either pembrolizumab TBP or salvage chemotherapy. Subsequent therapy was simulated for 18 weeks with radiographic assessment for response every 6 weeks. B. PFS2 in patients receiving pembrolizumab TBP, salvage chemotherapy, or a hypothetical PFS-optimized treatment (see Methods). ***, p < 0.001 (log-rank test). C. (B) for patients with target progression without nontarget progression. ***, p < 0.001 (log-rank test). D. (B) for patients with nontarget progression without target progression. ***, p < 0.001 (log-rank test). E. (B) for patients with simultaneous target and nontarget progression at radiographic assessment. ***, p < 0.001 (log-rank test). F. Best response, ORR, and DCR in patients receiving pembrolizumab TBP, salvage chemotherapy, or a hypothetical PFS-optimized treatment. Horizontal dashed lines indicate thresholds for PD and PR; purple diamonds represent individual lesions; colored bars indicate patients with responses and no nontarget progression; gray bars indicate patients with responses and simultaneous nontarget progression.

We hypothesized that carefully selecting patients to maintain on pembrolizumab TBP would produce better cohort-level outcomes than unconditionally selecting salvage chemotherapy. To investigate this, we simulated a hypothetical “PFS-optimized” regimen: whichever therapy expected to result in the longest PFS2 was provided to the patient at initial PD. It is worth nothing that this is currently impractical to implement in clinical practice given the lack of biomarkers guiding therapy selection in the pembrolizumab-refractory, PD-L1-high, driver alteration-free setting. Nevertheless, if salvage chemotherapy were optimal for cohort-level outcomes, we expected there to be no difference in PFS between the PFS-optimized regimen and the salvage chemotherapy regimen. Instead, we found that salvage chemotherapy significantly underperformed the PFS-optimized regimen in patients with nontarget progression, but not in patients with target-only progression (Figure 3B-E). Interestingly, while salvage chemotherapy underperformed the PFS-optimized regimen in DCR (67.9% vs. 81.1%), it produced a comparable ORR (32.3% vs. 32.1%), indicating that the primary benefit of the PFS-optimized regimen was through maintenance of stable disease as opposed to deepening of response.

### A subset of nontarget progressors may benefit from pembrolizumab beyond progression

Given the room to improve cohort-level PFS2 above salvage chemotherapy, we next attempted to identify the progressors who stood to benefit most from pembrolizumab TBP. When analyzing all progressors or target-only progressors, pembrolizumab TBP was inferior to salvage chemotherapy in PFS2 and control of tumor burden (Figure 3B-C, Figure 4A-B). However, pembrolizumab TBP and salvage chemotherapy produced indistinguishable PFS2 and control of tumor burden in patients with nontarget progression (Figure 3D-E, Figure 4C-D). As the PFS-optimized regimen was significantly better than both pembrolizumab TBP and salvage chemotherapy in this group, we inferred the existence of a subset of nontarget progressors who indeed benefited more from pembrolizumab TBP than salvage chemotherapy [7]. In these patients, moreover, pembrolizumab likely maintains SD as opposed to mediating a deeper reduction in tumor burden, as suggested by the discrepancy in ORR and DCR of salvage chemotherapy and PFS-optimized therapy (Figure 3F).

**Figure 4.**
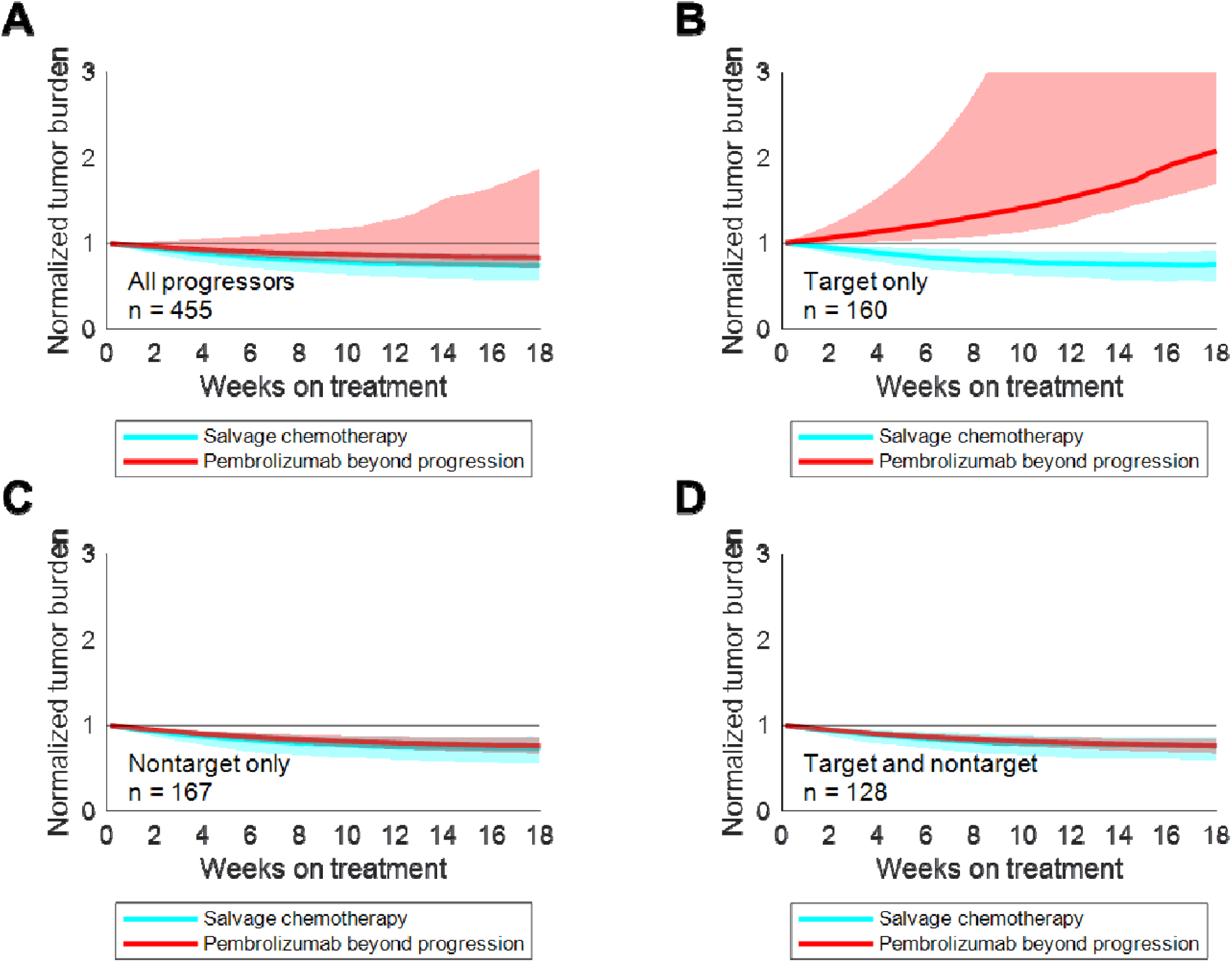
Pembrolizumab TBP can control tumor burden in nontarget progressors. A. Median tumor burden in patients receiving pembrolizumab TBP (red) or salvage chemotherapy (cyan). Shaded regions represent the interquartile range at each time point. B. (A) for patients with target progression without nontarget progression. C. (A) for patients with nontarget progression without target progression. D. (A) for patients with simultaneous target and nontarget progression.

### Nontarget progressors who benefit from pembrolizumab treatment beyond progression tend to have fewer lesions at baseline

Having identified nontarget progressors as the most likely to benefit from pembrolizumab TBP, we then tried to identify additional patient characteristics that would inform therapeutic selection. Nontarget progressors for whom pembrolizumab led to longer PFS2 were few in number (36 of 295; 12%) and tended to have fewer lesions at baseline (Figure 5A). However, they did not have greater baseline tumor burden nor a different anatomical lesion distribution (Figure 5B-C). We did not observe a difference in patterns of progression across anatomical sites at PD that would help select patients for pembrolizumab TBP (Figure 5D).

**Figure 5.**
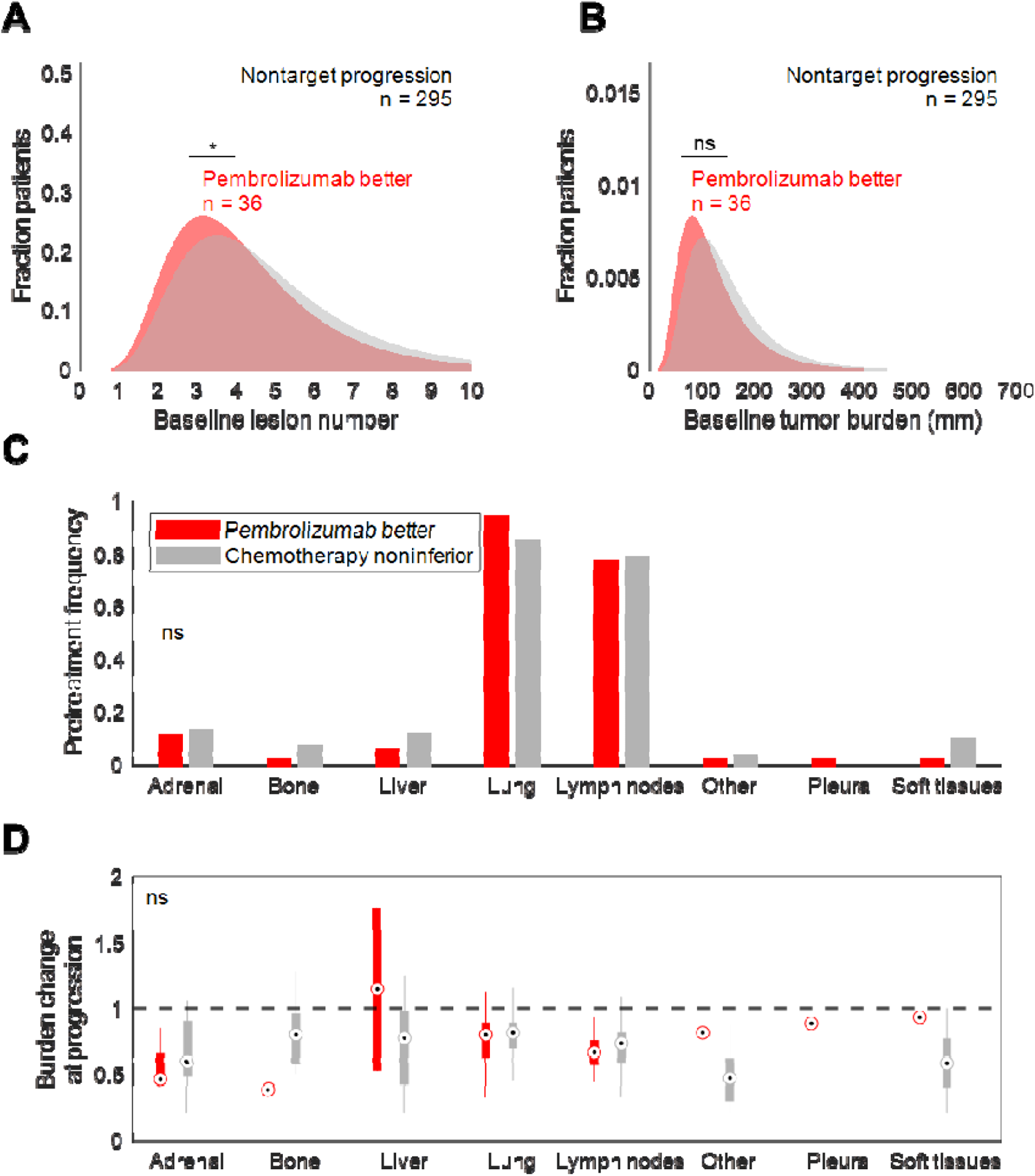
Nontarget progressors who benefit more from TBP have fewer lesions. A. Log normal distribution fit to baseline lesion number of nontarget progressors who experienced longer PFS on pembrolizumab TBP (red) or in whom salvage chemotherapy produced noninferior or better PFS (gray). *, p < 0.05 (unpaired Student’s t-test on raw data). B. (A) for baseline tumor burden. Ns, not significant (unpaired Student’s t-test on raw data). C. (A) by fraction of patients with baseline lesions in each anatomical site. Ns, not significant (Chi-squared test). D. (A) by tumor burden relative to baseline in each anatomical site. Ns, not significant (unpaired Student’s t-test).

### LLIA may improve response rates without extending progression-free survival

We next evaluated whether patients could benefit from combination therapy and, if so, how. Virtual patients were treated with combination therapy under either the PLIA or LLIA framework (Figure 6A, see Methods). LLIA but not PLIA led to a statistically significant improvement in PFS over chemotherapy alone (Figure 6B). ORR and DCR were also higher for LLIA (49.2%, 62.8%) than for PLIA (42.3%, 61.1%) and chemotherapy (29.7%, 57%). LLIA may represent a hitherto understudied mechanism through which drug synergy is derived.

**Figure 6.**
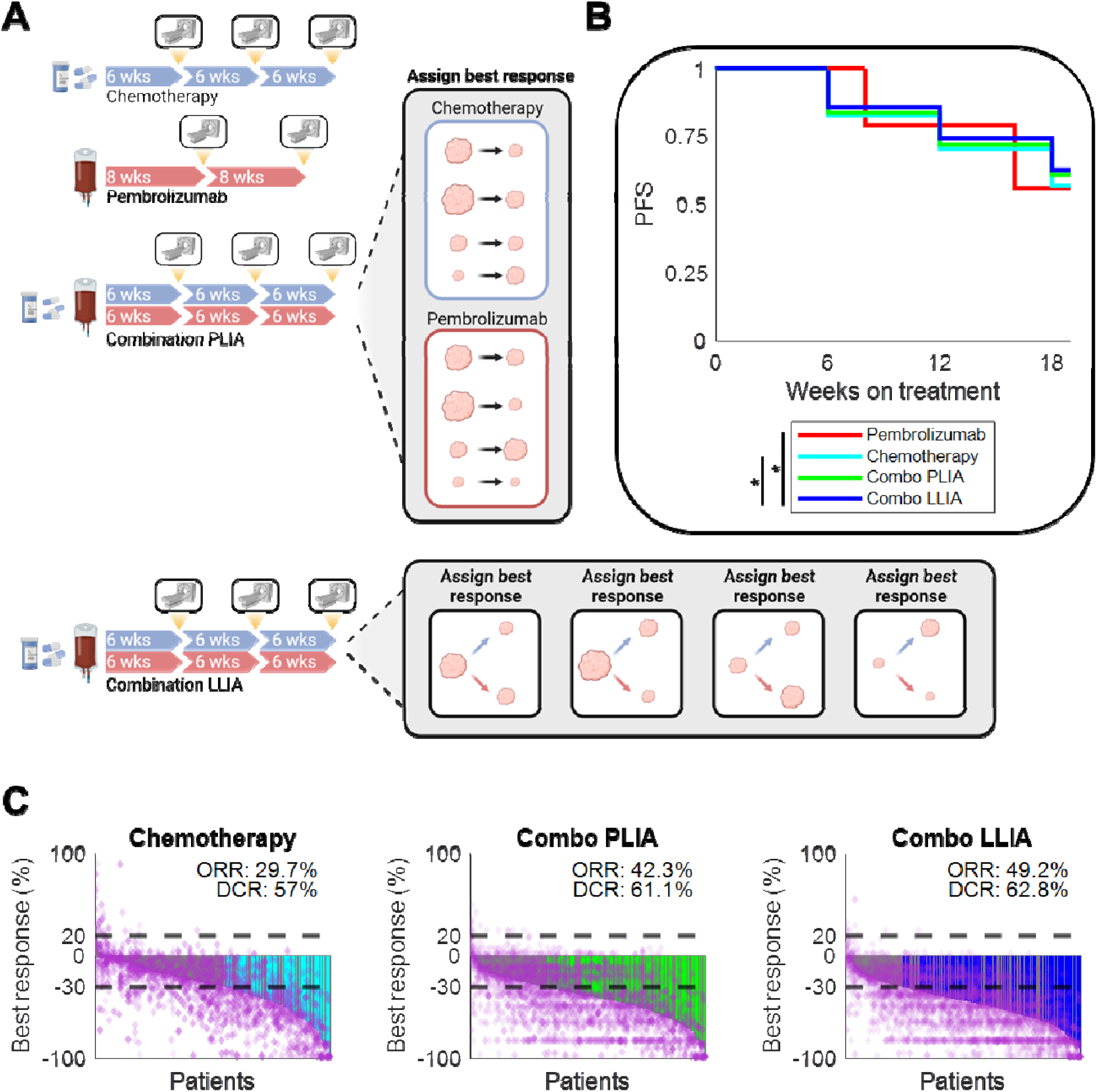
Combination therapies exhibiting LLIA could yield higher ORR and PFS than those exhibiting PLIA. A. Schematic of virtual clinical trial. Combination treatment with pembrolizumab and chemotherapy was simulated for 18 weeks with radiographic assessment for response every 6 weeks. PLIA and LLIA were evaluated separately as potential implementations of independent drug action. B. PFS2 in patients receiving pembrolizumab monotherapy, chemotherapy, PLIA combination therapy, or LLIA combination therapy. *, p < 0.05 (log-rank test). C. Best response, ORR, and DCR in patients receiving pembrolizumab monotherapy, chemotherapy, PLIA combination therapy, or LLIA combination therapy. Horizontal dashed lines indicate thresholds for PD and PR; purple diamonds represent individual lesions; colored bars indicate patients with responses and no nontarget progression; gray bars indicate patients with responses and simultaneous nontarget progression.

## DISCUSSION

Our study leveraged the intrapatient heterogeneity of response commonly observed under checkpoint blockade to explore the feasibility of treatment beyond progression with pembrolizumab in patients with PD-L1 TPS ≥ 50% [2], [23]. We found that patients whose target lesions progress under pembrolizumab are unlikely to derive benefit from pembrolizumab TBP. Conversely, those who had nontarget progression were most likely to benefit, perhaps because their target lesions were still shrinking; in particular, nontarget progressors with fewer lesions at baseline were the most likely to benefit from TBP over salvage chemotherapy. One possible mechanism underlying this observation could be that the absence of multiple small metastases evidences a degree of systemic tumor control by the immune system, whereas a highly disseminated pattern of tumor burden could indicate systemic immune failure. In practice, we expect it would be difficult to stratify based on baseline lesion number alone given the relatively small difference we observed.

Notably, while pembrolizumab plus pemetrexed and platinum chemotherapy is currently the recommended first-line therapy for previously untreated advanced NSCLC with PD-L1 TPS < 50% [1], [24], we did not have access to lesion growth dynamics data for patients treated with this regimen. We advocate for trial investigators to make lesion-level data available whenever feasible, as these data are instrumental to robust studies of tumor growth dynamics studies under diverse treatment regimens. Our own assessment of a pembrolizumab combination therapy found improvements in ORR under assumptions of LLIA compared to PLIA, despite their identical PFS curves. This suggests that PLIA as a descriptive framework could be improved by accounting for response rates in addition to PFS [11], [12]. Combination therapies that exhibit LLIA yield higher ORR than combination therapies that exhibit PLIA but are unidentifiable when assessed on PFS alone. While but one early example, we propose here LLIA as a putative mechanism by which drug synergy can be achieved. “Drug synergy” here differs slightly from the definition advocated by Palmer et al., as the benefit we observed above PLIA was not in PFS, but rather in response rate. Investigators leading future combination therapy studies might benefit from carefully assessing whether their chosen regimen is expected to display PLIA or LLIA.

Our simulations also yielded a modest increase in response rate to salvage chemotherapy after progression on pembrolizumab (32.3% vs. 29.7%), in agreement with previous reports that first-line pembrolizumab significantly enhances response rates to salvage chemotherapy compared to first-line chemotherapy [25], [26]. This may have been due to pembrolizumab and chemotherapy exerting differential efficacy in different anatomical sites, such that pembrolizumab “pre-treatment” shrunk tumors in anatomical sites that otherwise would have responded poorly to chemotherapy – i.e., one therapy covering the other’s bases with respect to biodistribution or site-specific efficacy. The differential biodistribution of chemotherapy, anti-PD-1 antibodies, and invigorated effector T cells remains underexamined in the setting of combination and sequential therapy.

One potential limitation of our study was the absence of simulated growth dynamics for new lesions, for which diameters were not reported in our source dataset. While negligible in the simulated monotherapy and combination therapy trials, it is possible that growth of these nontarget lesions in the sequential therapy trial might have caused some patients receiving subsequent therapy to reach PD more quickly. We considered this unlikely for two reasons: first, these patients by definition have tumor burden of at least 20% greater than baseline, causing a second increase of 20% to require substantial additional tumor mass; second, new metastases are likely to be smaller than extant lesions. Metastatic lesion measurements might have helped us assess patients via the immune RECIST (iRECIST) criteria and more accurately determine progression on pembrolizumab at radiographic assessments 4 – 8 weeks after initial unconfirmed PD [27]. Overall, our findings broadly aligned with those of Metro et al., who found pembrolizumab TBP to be beneficial for NSCLC patients with progression in ≤ 2 organ sites [7], and support the initiation of prospective studies of pembrolizumab TBP in patients with PD-L1-high, driver alteration-free NSCLC who experience nontarget progression.

## Data Availability

All data produced in the present study are available upon reasonable request to the authors.

## ABBREVIATIONS

PLIA: patient-level independent action
LLIA: lesion-level independent action

## AUTHOR CONTRIBUTIONS

Conceptualization, T.Q. and Y.C.; Methodology, T.Q.; Software, T.Q.; Validation, T.Q.; Formal Analysis, T.Q.; Investigation, T.Q.; Resources, Y.C.; Data Curation, T.Q.; Writing – Original Draft, T.Q.; Writing – Review & Editing – T.Q. and Y.C.; Visualization – T.Q.; Supervision – Y.C.; Project Administration – T.Q. and Y.C.; Funding Acquisition – Y.C.

## ACKNOWLEDGEMENTS

The authors thank Brian Topp, Kapil Mayawala, and Dinesh de Alwis for their helpful comments during study design. Figures were prepared in BioRender.

## ETHICS STATEMENT

This study was conducted using publicly available data:

- Study 1: Project DataSphere PDS UID: LungNo_Celgene_2007_108, link: https://data.projectdatasphere.org/projectdatasphere/html/content/108.
- Study 2: Nishino et al. (2021), link: https://doi.org/10.1200/PO.20.00478. The ethics oversight body that approved the collection of data associated with this study was not disclosed by the original authors.

The original studies on which these analyses were performed were given ethical approval by the institutional review boards at each participating center and conducted in accordance with the principles of the Declaration of Helsinki and Good Clinical Practice Guidelines of the International Conference on Harmonization. All patients provided written informed consent before study initiation. Of note, access to and permission to use the data associated with Study 1 requires completing an application for access via Project DataSphere’s website; this is generally granted within 7 business days. Such permission was granted for this study.

---

**Supplemental Figure 1.**
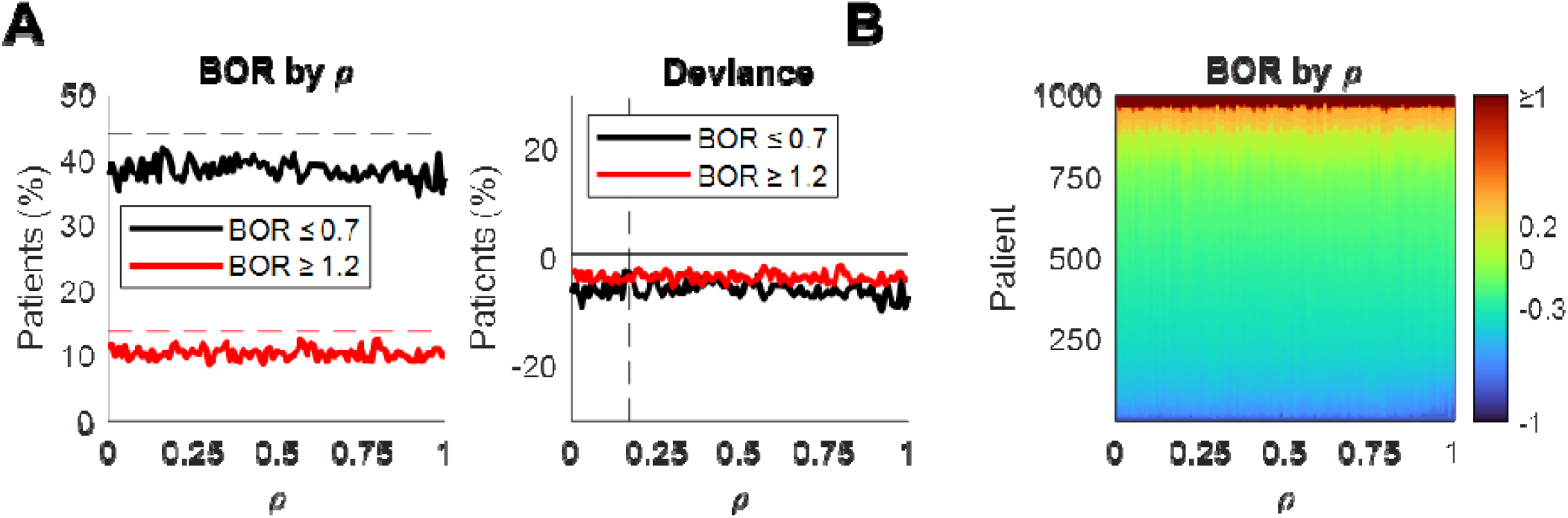
Responses to pembrolizumab were unaffected by variable inter-lesion correlation. A. Left, fraction of patients achieving CR/PR (black) or PD (red) at *ρ* values from 0 to 1. Horizontal dashed lines indicate fractions reported in [18] for pretreated NSCLC patients who received pembrolizumab. Right, deviance of fractions from reported values. The vertical dashed line indicates the *ρ* value (0.71) at which the sum of CR/PR deviance and PD deviance was minimized. B. Heatmap representation of BOR distributions at increasing levels of *ρ*.

**Supplemental Figure 2.**
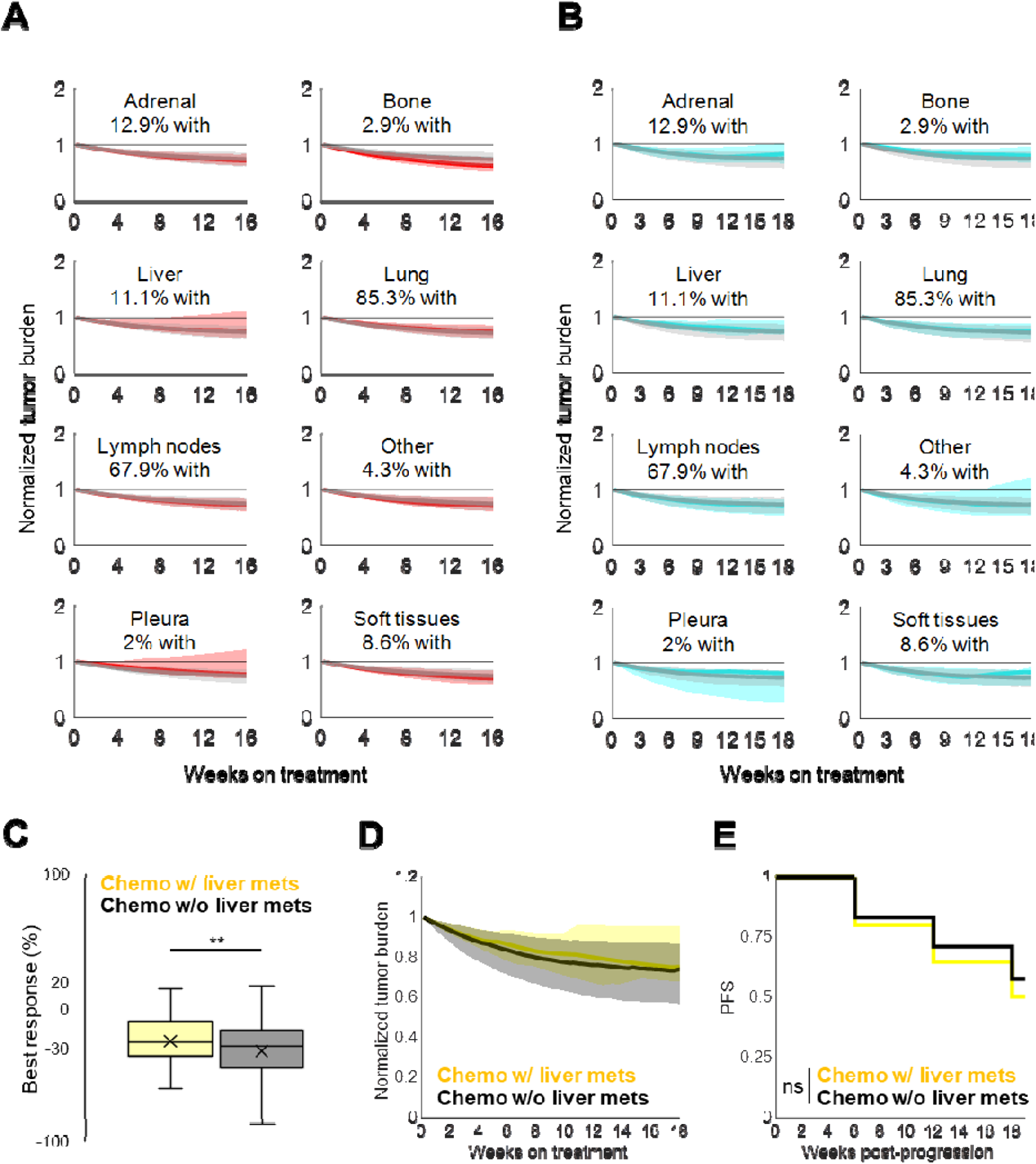
Lesion growth dynamics under virtual monotherapy vary across anatomical sites. A. Median tumor burden under pembrolizumab in patients with (red) or without (black) lesions in each anatomical site. Shaded regions represent the interquartile range at each time point. B. Median tumor burden under chemotherapy in patients with (cyan) or without (black) lesions in each anatomical site. Shaded regions represent the interquartile range at each time point. C. Best patient-level reduction in tumor burden under chemotherapy in patients with (yellow) and without (black) baseline liver metastases. **, p < 0.005 (unpaired Student’s t-test). D. Median tumor burden under chemotherapy in patients with (yellow) and without (black) baseline liver metastases. Shaded regions represent the interquartile range at each time point. E. PFS under chemotherapy in patients with (yellow) and without (black) baseline liver metastases. Ns, not significant (log-rank test).

